# COVID-19 VACCINE PERCEPTIONS AND DIFFERENCES BY SEX, AGE, AND EDUCATION: FINDINGS FROM A CROSS-SECTIONAL ASSESSMENT OF 1367 COMMUNITY ADULTS IN ONTARIO

**DOI:** 10.1101/2021.05.04.21256489

**Authors:** Sabrina K. Syan, Mahmood Gohari, Emily E. Levitt, Kyla Belisario, Jessica Gillard, Jane DeJesus, James MacKillop

**Affiliations:** Peter Boris Centre for Addictions Research, McMaster University & St. Joseph’s Healthcare Hamilton, Hamilton, Ontario, Canada

**Author notes:** <u>Corresponding Author:</u> Dr. James MacKillop., Peter Boris Centre for Addictions Research, St. Joseph’s Healthcare Hamilton, West 5th Campus/McMaster University, 100 West 5th Street, Hamilton, ON L8P 3R2, Canada. E-mail address (J. MacKillop).

**Keywords:** COVID-19, Vaccine, Attitudes, Hesitancy

## Abstract

**Background:** COVID-19 is a global pandemic and vaccination efforts may be impeded by vaccine hesitancy. The present study examined willingness to receive COVID-19 vaccine, the associated reasons for willingness/unwillingness, and vaccine safety perceptions in a cross-sectional assessment of community adults in Ontario.

**Methods:** 1367 individuals (60.3% female, M age = 38.6) completed an online assessment between January 15, 2021 and February 15, 2021. Perceptions were investigated in general and by age, sex and education.

**Results:** Overall, 82.8% sample reported they were willing to receive a COVID-19 vaccine and 17.2% reported they were unwilling. The three most common reasons for unwillingness were long-term side effects (65.5%), immediate side effects (60.5%), and lack of trust in the vaccine (55.2%). Vaccine willingness significantly differed by sex and education level, with female participants and those with less than a bachelor’s degree being more likely to report unwillingness. Perception of COVID-19 vaccine safety was significantly lower (−10.7%) than vaccines in general and differed by age, sex and education, with females, older adults, and individuals with less than a bachelor’s degree reporting lower perceived COVID-19 vaccine safety.

**Conclusion:** In this sample of community adults, under one in five individuals was unwilling to receive a COVID-19 vaccine, but with higher rates in population subgroups. Targeting public health messaging to females and individuals with less than Bachelor’s degree, and addressing concerns about long-term and immediate side effects may increase vaccine uptake.

## INTRODUCTION

The novel coronavirus disease (COVID-19) caused by severe acute respiratory syndrome coronavirus 2 (SARS-CoV-2) was declared a global pandemic by the World Health Organization in March 2020. To date, approximately 124 million individuals globally have been infected with COVID-19 and approximately 2.7 million individuals have died(*COVID-19 Map - Johns Hopkins Coronavirus Resource Center*, n.d.). Global efforts to mitigate the spread of COVID-19 included travel restrictions, international border closures, and strong public health measures such as physical distancing measures and mask mandates. Complementing these strategies, vaccination against COVID-19 substantially mitigates severity of COVID-19. To date, four vaccinations to protect against COVID-19 infection and symptoms have been approved by Health Canada, three of which are currently being offered to Canadians.

As a mass inoculation program begins, one potential impediment is vaccine hesitancy (i.e., unwillingness to receive the vaccine). The goal of this study was to clarify the proportion of the population likely to decline a COVID-19 vaccine and the associated reasons. To do so, we examined the prevalence of vaccine willingness/unwillingness and the reasons associated with each perspective in a cross-sectional assessment of general community adults in Ontario. In addition to general perceptions, we examined differences on the basis of sex, age, and education; and perceptions of safety of vaccines in general compared to COVID-19 vaccines. Clarifying reasons for vaccine unwillingness and subgroup differences have the potential to inform public health strategies to address specific concerns and improve vaccine education.

## METHODS

### Participants and Study Design

This study capitalized on an ongoing longitudinal surveillance study investigating health-related behaviors among community adults in Southern Ontario. Individuals in this larger community-based study were recruited through advertisements (e.g., social media, print and online advertising) from 2016-2018 in Hamilton, ON for an in-person assessment and have received periodic follow-up assessments. Original eligibility criteria were that participants were 18-65 years of age, have at least a ninth-grade education (for literacy), had no extant terminal illness, and indicated a willingness to consider participation in future research studies. The exclusion criteria for this study were purposely minimal to maximize enrollment. Individuals enrolled in the study were non-clinical, community dwelling, healthy adults that were interested in participating in health research studies. As part of this ongoing study, information related to vaccine administration and hesitancy was collected in the most recent data collection wave, from January 15, 2021 to February 15, 2021. The sample comprised 1367 individuals from the original a longitudinal cohort of 1502 (Table 1). Participants reflected middle-aged adults with greater representation of females compared to males (60.6%). Data were collected via Research Electronic Data Capture (REDCap) software(Harris et al., 2009) and participants received a $40 gift card. The study was approved by the Hamilton Integrated Research Ethics Board (Protocol # 4699) and complied with the Helsinki Declaration.

**Table 1:** Participant demographics (n = 1367)

| Demographic Variable | Mean (SD)/ % |
| --- | --- |
| *Age* | 38.6 (14) |
| *Sex* |  |
| Male | 543 (39.7%) |
| Female | 824 (60.3%) |
| *Education* |  |
| < Bachelor's Degree | 472 (34.5%) |
| Bachelor's Degree | 660 (48.3%) |
| >Bachelor's Degree | 235 (17.2%) |
| *Race* |  |
| White | 1100 (80.5%) |
| Black | 18 (1.3%) |
| Asian | 152 (11.1%) |
| First Nations/Inuit/Metis | 13 (1.0%) |
| Pacific Islander | 5 (0.4%) |
| More than one population group | 51 (3.7%) |
| Other | 28 (2.0%) |

### Assessment

The assessment is provided in Supplementary Information and comprised purpose-built questions to ascertain willingness to receive the COVID-19 vaccine and examined potential reasons for either receiving or declining the vaccine. Questions regarding the perception of COVID-19 vaccine safety (i.e., “how safe do you believe the COVID-19 vaccines are?”) and general vaccine safety (i.e. “how safe do you believe vaccines are in general?”) were asked using a single item visual analogue scale ranging from 0 to 100 with anchors of very unsafe (0) to very safe (100).

### Data Analysis

Five quality control questions with unambiguously correct answers (e.g., “in response to this question, please choose option “nearly every day”) were included and participants were excluded for 2+ incorrect responses or if they did not complete the entire survey. Analyses were restricted to unvaccinated individuals. Age and educations subgroups were constructed to map on to commonly used distinctions while approximately balancing sample sizes. Categorical variables were compared across groups using a χ^2^ test. Independent sample T-tests were used to compare continuous variables across two groups. Analyses of variance was used to compare continuous variables across three groups. All statistical analysis was completed using SAS Software and the criterion for statistical significance was *p*<0.05.

## RESULTS

### Vaccine Willingness

A small subset reported having received at least one dose (5.1%, *n* = 70) at the time of participation and were not examined further. More than 4 in 5 participants had a favourable perspective on the vaccine (82.8%) (Table 2). Reasons for willingness are in Figure 1, with prevention of transmission (91%) and protection from contracting COVID-19 (90%) being the most endorsed. For the 17.8% reporting unwillingness, the reasons for unwillingness are in Figure 2, with long-term side effects (65.5%), immediate side effects (60.5%), and a lack of trust in the vaccine itself (55.2%) being the three most common reasons for not wanting to receive the vaccination.

**Table 2:** COVID-19 vaccination willingness among unvaccinated participants in general and by subgroup (n=1297)

| Demographic Subgroup | Unwilling (No) | Willing (Yes) | $\chi^2$ | P |
| --- | --- | --- | --- | --- |
| **Total Sample** | 223 (17.2%) | 1074 (82.8%) |  |  |
| **Sex** |  |  | 9.71 | 0.002 |
| Male | 69 (13.1%) | 457 (86.9%) |  |  |
| Female | 154 (19.9%) | 617 (80.1%) |  |  |
| **Age (years)** |  |  | 1.72 | 0.423 |
| <30 | 80 (15.5%) | 436 (84.5%) |  |  |
| 30-49 | 82 (18.4%) | 364 (81.6%) |  |  |
| 50+ | 61 (1.82%) | 274 (81.8%) |  |  |
| **Education** |  |  | 63.7 | <0.001 |
| < Bachelors Degree | 155 (25.9%) | 443 (74.1%) |  |  |
| Bachelors Degree | 57 (11.7%) | 430 (88.3%) |  |  |
| >Bachelors Degree | 11 (5.2%) | 201 (94.8%) |  |  |

Note: The frequencies above were calculated by $\chi^2$ between the percent of individuals that indicated that they were willing to receive the vaccination (Yes) or unwilling to receive the vaccination (No).

**Figure 1:**
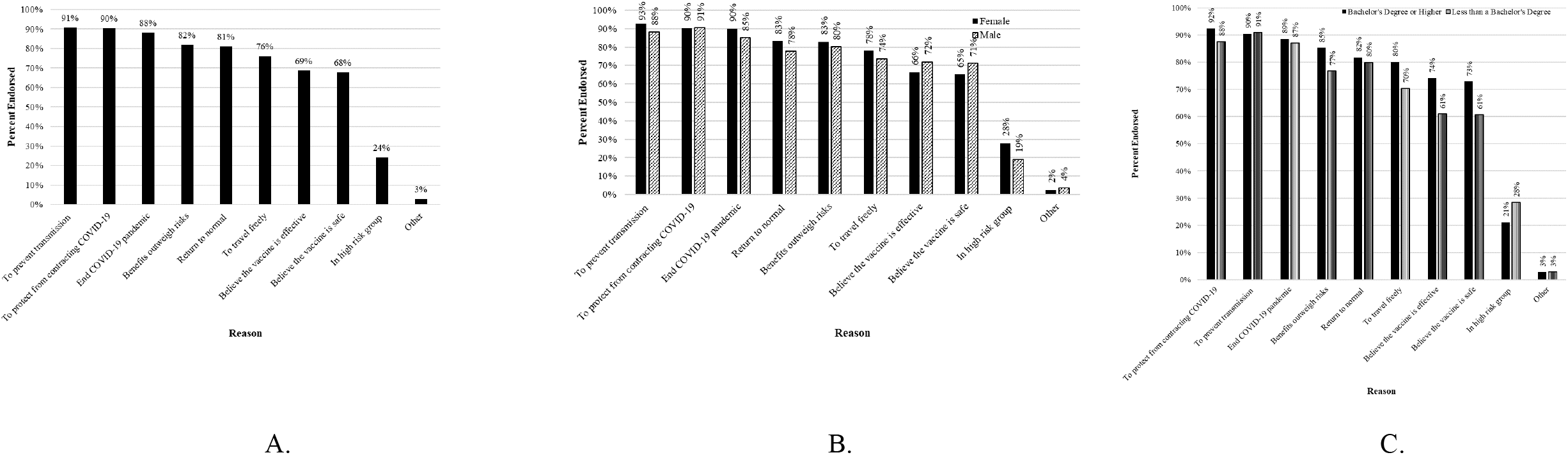
Reasons for receiving the COVID-19 vaccine reported by participants that indicated vaccine willingness. **Panel A:** Overall reasons for receiving the COVID-19 vaccine. **Panel B**: Reasons for receiving the COVID-19 vaccine reported between male and female participants. **Panel C**: Reasons for receiving the COVID-19 vaccine reported between participants with less than a Bachelor’s degree or a Bachelor’s degree or higher.

**Figure 2:**
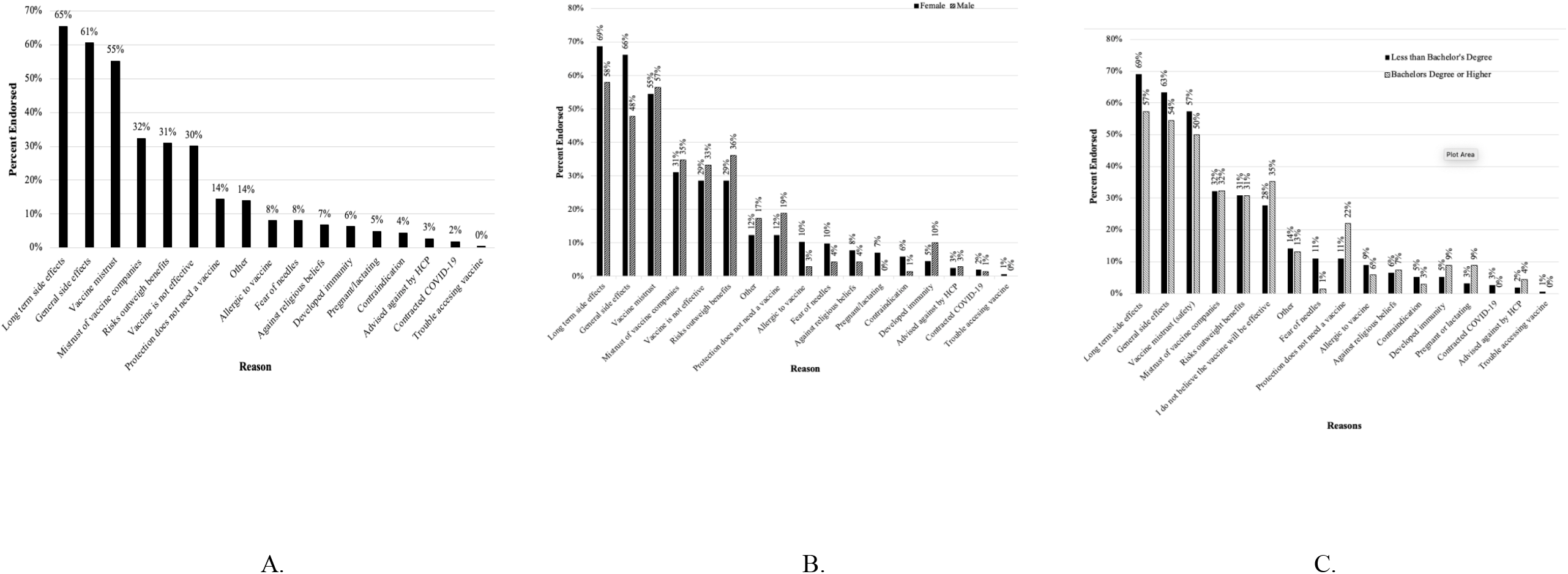
Reasons for not wanting to receive the COVID-19 vaccine reported by participants that indicated vaccine unwillingness. **Panel A**: Overall reasons for not wanting to receive the COVID-19 vaccine. **Panel B**: Reasons for not wanting to receive the COVID-19 vaccine across sex. **Panel C**: Reasons for not wanting to receive the COVID-19 vaccine reported between participants less than a Bachelor’s degree or a Bachelor’s degree or higher

Willingness to receive the COVID-19 vaccine differed by sex and education, but not age (Table 2). Males reported modestly higher willingness than females (87% vs. 81.3%). With regards to education, individuals with lower than a bachelor’s degree were also more likely to refuse a COVID-19 vaccination; 25% of individuals with less than a bachelor’s degree education indicated that they were not willing to receive a COVID-19 vaccine, as compared to 11% with a bachelor’s degree level education and 4.7% with at least a graduate level education (Table 2).

Differences in reasons for willingness by sex and education are in Figures 1 and 2; female participants and those with higher education reported greater endorsement of wanting to prevent transmission and protection from contracting COVID-19. Differences in reasons for unwillingness by sex and education are in Figure 2. In this case, female participants and those with less than a bachelor’s degree reported that they worried about long term and immediate vaccine side effects than male participants and participants with a bachelor’s degree or higher.

### Vaccine Safety

With regards to vaccine safety, the COVID-19 vaccine was rated 10.7% less safe than a general vaccine; participants endorsed significantly greater perception of general vaccine safety (84.2) as compared to COVID-19 vaccine safety (73.5). Subgroup differences were present based on sex, age, and education (Table 3). Male participants endorsed higher vaccine safety perception for general vaccines and the COVID-19 vaccines, compared to female participants. Individuals under 30 years of age endorsed a significantly higher perception of both general and COVID-19 vaccine safety as compared to individuals in the oldest age bracket, over 50 years of age (p<0.05). General or COVID-19 vaccine safety perceptions did not differ significantly between the under 30- and 30-49-year age brackets. Individuals with less than a bachelor’s degree indicated lower perception of general and COVID-19 vaccine safety than both higher education groups (*p*s<0.05) and individuals with greater than a bachelor’s degree (*p*<0.05). There was not a significant difference in perception of general or COVID-19 vaccination safety between individuals with a bachelor’s degree and those with greater than a bachelor’s degree.

**Table 3:** Perception of general and COVID-19 vaccine safety among participants (n=1367). Note that COVID-19 vaccines were not differentiated by specific product.

|  | General Vaccine Safety<br>Perception (0-100) |  | COVID-19 Vaccine Safety<br>Perception (0-100) |  |
| --- | --- | --- | --- | --- |
|  | Mean<br>(SEM) | *p* | Mean<br>(SEM) | *p* |
| **Total** | 84.2 (0.53) |  | 73.5 (0.69) |  |
| **Sex** |  | <0.001 |  | 0.001 |
| Male | 86.5 (0.75) |  | 76.6 (1.04) |  |
| Female | 82.6 (0.72) |  | 71.5 (0.91) |  |
| **Age (years)** |  | <0.001 |  | 0.021 |
| <30 | 86.6 (0.72) |  | 75.8 (1.02) |  |
| 30-49 | 82.8 (0.96) |  | 72.6 (1.21) |  |
| 50+ | 82.3 (1.11) |  | 71.3 (1.41) |  |
| **Education** |  | <0.001 |  | <0.001 |
| < Bachelors Degree | 80.2 (0.89) |  | 66.8 (1.13) |  |
| Bachelors Degree | 87.1 (0.74) |  | 78.5 (0.95) |  |
| > Bachelors Degree | 88.7 (0.94) |  | 81.2 (1.32) |  |

## DISCUSSION

This study evaluated willingness to receive the COVID-19 vaccination and perceptions of vaccine safety of the COVID-19 vaccination in a community-based sample of adults from Southern Ontario. Results indicate that although a large proportion of the sample was willing to receive the vaccination, more than 4 in 5, a sizable fraction of the population (17.2%) was not. Individuals who are not willing to receive the vaccination indicated that concerns regarding immediate and long-term vaccine side effects and a lack of trust in the vaccine itself are their main reasons for not wanting to receive the vaccination. Vaccine willingness increased with level of education and significantly differed between male and female participants, with female participants reporting that they are less willing to be vaccinated.

Participants reported that they considered the COVID-19 vaccine 10.7% less safe than they viewed general vaccine safety. This may be due to the speed with which the COVID-19 vaccination was created, and expedited approval times associated with the vaccinations as compared to general vaccination development process which may take a decade or longer. Furthermore, the evolving narrative surrounding information regarding COVID-19, COVID-19 variants and the efficacy of vaccinations against potential variants may also be a contributing factor to vaccine hesitancy. Specific to female participants, concerns regarding vaccinations and fertility may play a role in making female participants less willing to receive the COVID-19 vaccine and may suggest that they believe it is less safe. It should be noted that 7% of women reported that their unwillingness to receive the vaccination was due to pregnancy or lactation. Recent evidence regarding the safety and efficacy of the COVID-19 vaccinations in pregnant and lactating women and conferred benefits to newborns as a result are emerging(Gray et al., 2021), but would not have been available at the time of the assessment

Perceptions of general and COVID-19 vaccine safety both decreased with age and increased with education. Perceptions of vaccine safety (but not willingness) differed by age; although older adults (50 years of age or older) reported that they were still willing to receive the vaccination. Vaccine willingness did not differ by age, however, did differ by education - with individuals with less than a Bachelor’s degree indicating that they 13.1% less willing as individuals with greater than a Bachelor’s degree to receive the vaccination. This is consistent with recent findings from the United States that individuals with higher education are significantly more likely to get a COVID-19 vaccination and to believe in the effectiveness and safety of the vaccine(*Education Is Now a Bigger Factor than Race in Desire for COVID-19 Vaccine*, n.d.). Education is also associated with greater engagement in pro-health behaviours(Park et al., 2018; Zajacova & Lawrence, 2018) such as vaccinations and lower delay discounting(Daugherty & Brase, 2010), a measure of impulsivity that refers to the decline in value of a reward based on the delay to its receipt that has been linked to numerous health behaviors, including vaccine uptake(Bradford, 2010; Chapman & Coups, 1999).

The results of this manuscript highlight several implications for public health messaging to maximize vaccine uptake. The most common concerns highlighted by individuals in our study that were unwilling to receive the vaccine included concerns regarding long term and more general vaccine side effects and general vaccine mistrust. Effective public health messaging to combat vaccine hesitancy should focus on providing information regarding the immediate and long-term vaccine side effects in an effort to improve vaccine willingness and perceptions of vaccine safety. Further, results highlight that female participants and those with less than a bachelor’s level education endorsed greater concerns with regards to long-term and general vaccine willingness as compared to male or more educated participants and may need to be the focus of enhanced public health messaging. Targeting public health messaging to these subpopulations may increase vaccine willingness.

This study should be considered in the context of its strengths and limitations. Amongst its strengths, it systematically assessed vaccine willingness, reasons, and safety, and provides timely information that may guide public health efforts to decrease hesitancy and increase vaccine uptake. The sample size was relatively large which allowed for high statistical power and the sample was relatively representative of community adults in Canada. For example, approximately 65% of our sample completed at least some postsecondary education, which is consistent with the national average of 68%. Further, the median age, racial demographics and income reported within the study are reflective of the general population in Hamilton, ON, thus, the results of this study are generalizable to the wider population of the region. With regards to limitations, our study population was primarily comprised of individuals of European ancestry and thus lacks racial diversity. As such, we were not able to comment on willingness to receive the COVID-19 vaccination or safety perceptions within specific racial groups and may be less generalizable to highly racially diverse catchment areas. Similarly, the sample was recruited from an urban/suburban catchment area and thus lacks rural representation.

Taken together, in this large community-based sample, the large majority of individuals reported being willing to receive the COVID-19 vaccination, but a notable fraction portion was not, principally due to concerns regarding vaccine side effects and a lack of trust in the vaccine itself. This suggests that public health messaging to combat vaccine hesitancy should focus on the safety profile of the approved COVID-19 vaccinations and consider targeting the segments of the population reporting the greatest unwillingness to vaccination.

## Data Availability

NA

