## Supplementary Tables for "COVID-19 VACCINE PERCEPTIONS AND DIFFERENCES BY SEX, AGE, AND EDUCATION: FINDINGS FROM A CROSS-SECTIONAL ASSESSMENT OF 1367 COMMUNITY ADULTS IN ONTARIO"

Supplementary Materials

Table S1: Assessment question to ascertain vaccine willingness

| Selection/Option | Question |
|---|---|
| 2 | Yes, I have already taken the vaccine |
| 1 | Yes, I will take the vaccine |
| 0 | No, I will not take the vaccine |

Table S2: Questions to clarify reasons for vaccine willingness (affirmative responses)

| Item | Question |
|---|---|
| 1 | To protect myself from contracting COVID-19 |
| 2 | To prevent transmission to my family, friends, or other contacts |
| 3 | I am in a high-risk group (e.g., elderly, immunocompromised, essential worker, etc.) |
| 4 | I believe the vaccine is safe |
| 5 | I believe the vaccine is effective |
| 6 | I believe the benefits outweigh the risks |
| 7 | To return to "normal" activities (e.g., no masks, no distancing, etc.) |
| 8 | To travel more freely |
| 9 | To help end the COVID-19 pandemic |
| 10 | Other |

Table S3: Questions to clarify reasons for vaccine unwillingness (negative responses)

| Item | Question |
|---|---|
| 1 | I have been advised by a healthcare provider not to get the vaccine |
| 2 | I have allergies or a history of allergies with vaccines |
| 3 | I have a previous symptom or condition that makes treatment risky (e.g., vaccine contraindication) |
| 4 | I am pregnant or lactating |
| 5 | I have a fear of needles |
| 6 | I do not trust that the vaccine is safe |
| 7 | I am worried about the side effects of the vaccine |
| 8 | I am worried about the long-term effects of the vaccine |
| 9 | I do not believe the vaccine will be effective |
| 10 | I believe the risks outweigh the benefits |
| 11 | It goes against my personal or religious beliefs (e.g., Contains objectionable ingredients, not kosher/halal/vegan/cruelty free) |
| 12 | I do not trust the companies and/or governments providing the vaccine |
| 13 | I have or will have trouble accessing the vaccine (e.g., mobility issues or travel issues) |
| 14 | I have already contracted COVID-19 |
| 15 | I believe I have already developed immunity |
| 16 | I believe protection against COVID-19 does not require a vaccine |

| | |
|---|---|
| 17 | Other |

Table S4: Assessment questions that used a visual analogue scale

| **Question** | **Visual Analog Scale: 0 (Very Unsafe) – 100 (Very Safe)** |
|---|---|
| Please rate how safe you believe vaccines are in general | 0-100 |
| Please rate how safe you believe the COVID-19 vaccines are | 0-100 |
